# Traumatic brain injury and Alzheimer’s Disease biomarkers: A systematic review of findings from amyloid and tau positron emission tomography (PET)

**DOI:** 10.1101/2023.11.30.23298528

**Authors:** Kaitlyn M. Dybing, Cecelia J. Vetter, Desarae A. Dempsey, Soumilee Chaudhuri, Andrew J. Saykin, Shannon L. Risacher

**Affiliations:** Department of Radiology and Imaging Sciences, Indiana University School of Medicine, Indianapolis, IN, 46202, USA; Indiana Alzheimer’s Disease Research Center, Indiana University School of Medicine, Indianapolis, IN, 46202, USA; Stark Neurosciences Research Institute, Indiana University School of Medicine, Indianapolis, IN, 46202, USA; Department of Neurology, Indiana University School of Medicine, Indianapolis, IN, 46202, USA; Ruth Lilly Medical Library, Indiana University School of Medicine, Indianapolis, IN, 46202, USA; Department of Psychiatry, Indiana University School of Medicine, Indianapolis, IN, 46202, USA

## Abstract

Traumatic brain injury (TBI) has been discussed as a risk factor for Alzheimer’s disease (AD) due to its association with dementia risk and earlier cognitive symptom onset. However, the mechanisms behind this relationship are unclear. Some studies have suggested TBI may increase pathological protein deposition in an AD-like pattern; others have failed to find such associations. This review covers literature that uses positron emission tomography (PET) of amyloid-β and/or tau to examine subjects with history of TBI who are at risk for AD due to advanced age. A comprehensive literature search was conducted on January 9, 2023, and 24 resulting citations met inclusion criteria. Common methodological concerns included small samples, limited clinical detail about subjects’ TBI, recall bias due to reliance on self-reported TBI, and an inability to establish causation. For both amyloid and tau, results were widespread but inconsistent. The regions which showed the most compelling evidence for increased amyloid deposition were the cingulate gyrus, cuneus/precuneus, and parietal lobe. Evidence for increased tau was strongest in the medial temporal lobe, entorhinal cortex, precuneus, and frontal, temporal, parietal, and occipital lobes. However, conflicting findings across most regions of interest in both amyloid- and tau-PET studies indicate the critical need for future work in expanded samples and with greater clinical detail to offer a clearer picture of the relationship between TBI and protein deposition in older subjects at risk for AD.

## Introduction

Alzheimer’s disease (AD) is the most common form of dementia, with approximately 6.7 million Americans currently living with the disease and this number expected to grow over the coming decades as the population ages.^1^ Worldwide, it is estimated that by 2050, 152 million people will be living with AD.^2^ There are presently no cures for AD, though recent disease-modifying therapies have become available.^3^ There are two main forms of AD, classified based on age of onset: late-onset AD (onset aged 65 years or older) and early-onset AD (onset before the age of 65).^4^ The late-onset form of AD is the most common, representing approximately 95% of all AD cases.^5^ Generally, AD is clinically associated with progressive memory decline and cognitive impairment, and is pathologically defined by the presence of neurodegeneration and cortical accumulation of β-amyloid as amyloid plaques and hyperphosphorylated tau as neurofibrillary tangles (NFTs).^6,7^ A framework for describing AD as a function of the most widely used biomarkers is the A/T/N (amyloid/tau/neurodegeneration) classification system, which encompasses biomarkers of β-amyloid, tau, and neurodegeneration and/or neuronal injury.^8^ Recently proposed updates to this framework explore the utility of additional biomarker categories, including inflammation/astrocytic activation (“I”), vascular brain injury (“V), and α-synuclein (“S”), to better incorporate comorbid pathologies in suspected AD patients.

Amyloid and tau accumulate in characteristic patterns in typical AD. Amyloid pathology classically deposits in five stages or phases; this process can begin decades before the onset of cognitive symptoms in patients who will go on to develop dementia.^9,10^ Stage 1 is characterized by amyloid deposition in the frontal, temporal, parietal, or occipital neocortex. The allocortex (insula, entorhinal cortex, and CA1 of the hippocampus) is also positive for amyloid deposition in Stage 2.^9^ Stage 3 involves amyloid depositions in the diencephalon (thalamus and hypothalamus), striatum (caudate and putamen), and basal forebrain.^9^ Stage 4 typically sees amyloid deposition in various brainstem nuclei, including the substantia nigra, red nucleus, central gray, superior and inferior colliculi, inferior olivary nucleus, and intermediate reticular zone.^9^ Finally, Stage 5 is characterized by amyloid deposits in the cerebellum as well as other brainstem nuclei (pontine nuclei, locus coeruleus, parabrachial nuclei, reticulo-tegmental nucleus, dorsal tegmental nucleus, and raphe nuclei).^9^

Tau deposits in six stages known as Braak stages.^11^ In Stage 1, tau is predominantly located in the transentorhinal region.^11^ In Stage 2, lesions extend into the hippocampus and/or entorhinal cortex.^11^ In Stage 3, tau deposits are found in the fusiform, parahippocampal, and/or lingual gyri.^11^ Patients typically begin to show cognitive symptoms in approximately Braak Stage 4; at this stage, tau is present in all regions of Stages 1-3 but is also found in the inferior temporal gyrus and insula.^11^ Stage 5 is defined with widespread tau deposition in regions that include those in all previous stages, as well as the peristriate area in the occipital lobe, as well as other frontal, superolateral, and occipital directions.^11^ In Stage 6, the final defined stage, tau pathology is found in all stages seen in Stages 1-5, both secondary and primary neocortical areas, as well as the striate area of the occipital lobe.^11^

There are a variety of well-known risk factors for AD, the most well-characterized of which include advanced age, family history, and genetic variations (such as one or more *APOE* ε4 allele(s)).^12^ Traumatic brain injury (TBI), also called concussion or head injury, is another commonly-cited risk factor for dementia. TBIs are extremely common: over a quarter (28.9%) of American adults report experiencing at least one in their lifetime.^13^ TBIs occur even more frequently amongst athletes who play contact sports as well as military servicemembers.^14,15^ This high frequency means there is an urgent need to better understand the long-term consequences of TBI, including biological pathways and consequences related to dementia, as TBI affects a significant number of people across a wide array of demographics.

TBI has been studied as a risk factor for neurodegenerative disease in the context of the well-documented relationship between repetitive TBI and chronic traumatic encephalopathy (CTE), a neurodegenerative disease identified frequently in American football players and found in many other populations with high rates of repetitive TBI.^16-19^ Similarly, interest in TBI as a risk factor for AD has been growing with the reports of a link between TBI and earlier onset of AD.^20-23^ However, there is a lack of consensus in the literature, as some studies have failed to find such an association.^24,25^ As a consequence of these disparate findings, less attention has been directed towards understanding the biological underpinnings of the relationship between TBI and risk for AD. However, developing technologies, including the widespread adaptation of positron emission tomography (PET) as a tool for in-vivo detection and quantification of pathological protein buildup, have better enabled the field to analyze the spatial and temporal dynamics of protein accumulation in neurodegenerative diseases in the context of risk factors such as TBI.

Positron emission tomography (PET) is a radiological technique that can be used to visualize the concentrations and distributions of radiolabeled molecules across the body *in vivo*.^26,27^ It is widely used in dementia research for the purpose of detection and quantification of pathological changes associated with amyloid-β (Aβ) and tau, as well as other molecules in the brain. PET uses a small amount of a radioactive tracer that binds to a specific protein target such as Aβ or tau.^28^ The tracer emits positrons as a function of the radioactive decay process, and these positrons are detected by the scanner and computationally mapped back to their place of origin, enabling visualization of both the relative quantity and spatial positioning of protein buildup in the brain.^28^ There are a multitude of different tracers that have been developed to label Aβ and tau; we will briefly discuss those used in the studies cited in this review. The Aβ tracers include Pittsburgh Compound B ([^11^C]-PiB), Florbetapir ([^18^F]AV-45), Florbetaben ([^18^F]FBB), ^18^F-FPYBF-2, and Flutafuranol ([^18^F]-NAV4694), and the tau tracers include Flortaucipir ([^18^F]AV-1451) and Florquinitau ([^18^F]-MK-6240).

Pittsburgh Compound B ([^11^C]-PiB) is a thioflavin-T analog and was the first specific PET tracer for Aβ used in human subjects.^29^ It binds with high specificity and sensitivity to fibrillar Aβ.^30,31^ One of the most commonly utilized Aβ tracers, [^11^C]-PiB has been validated in numerous studies investigating AD.^32-34^ Compared to healthy controls, AD patients typically show increased retention in frontal, parietal, temporal, and occipital cortices, and the striatum, but relatively minimal signal in subcortical white matter, the pons, and cerebellum.^29,32^ [^11^C]-PiB is known to bind nonspecifically to white matter due to its lipophilic properties.^35^ It has proven utility in distinguishing between multiple types of dementia, including AD, frontotemporal lobar degeneration (FTLD), cerebral amyloid angiopathy (CAA), Parkinson’s disease (PD), and HIV dementia.^33,35-37^ However, a significant clinical limitation is its short radionucleotide half-life of only approximately 20 minutes.^38^

Florbetapir, also known as [^18^F]AV-45 or Amyvid, is a PET tracer which binds with high specificity to β-amyloid plaques.^39,40^ The use of radioactive ^18^F, which has a half-life of 110 minutes, makes Florbetapir advantageous compared to tracers like [^11^C]-PiB which has a shorter half-life.^39^ Florbetapir binding patterns observed in AD patients are similar to those seen in studies using [^11^C]-PiB.^29,32,39^ In patients with AD compared to controls, greater Florbetapir signal is seen in areas with high levels of pathologic amyloid deposition, such as the frontal and temporal cortices, anterior cingulate, and precuneus.^39,41^ However, Florbetapir shows nonspecific binding in cortical white matter.^42,43^ The high sensitivity and specificity of Florbetapir for β-amyloid plaques has been confirmed by multiple studies showing a strong correlation between pathologically-stained Aβ plaques and Florbetapir binding.^40,44,45^ Florbetapir can also be used to identify different varieties of myocardial amyloid, and may be useful in distinguishing between AD and frontotemporal dementia (FTD).^46,47^

Florbetaben, also called Neuraceq, [^18^F]-BAY94-9172 or [^18^F]FBB, is a PET tracer which binds with high sensitivity and specificity to β-amyloid plaques.^48,49^ Patients with AD typically show retention of Florbetaben in the frontal cortex, posterior cingulate gyrus, precuneus, and lateral temporal and parietal cortical areas.^48-50^ Similar to Florbetapir, Florbetaben exhibits relatively high off-target uptake in the white matter.^42,43,48,50^ Interestingly, Florbetaben shows a lack of sensitivity in the hippocampus and parahippocampal gyrus.^49^ However, its ability to discriminate between AD patients and healthy controls can be improved via partial volume correction.^51^ Florbetaben has been studied for use in asymptomatic patients to detect prodromal AD, and has also demonstrated utility in distinguishing AD patients from patients with FTLD.^48,50,52^

[^18^F]-FPYBF-2 is a newer PET tracer which targets β-amyloid plaques.^53,54^ It has been reported to have higher binding affinity for Aβ than [^11^C]-PiB, but has similar uptake patterns to [^11^C]-PiB and shows very little off-target binding.^53,55^ In patients with AD, [^18^F]-FPYBF-2 retention is seen in both gray and white matter, while in healthy controls uptake is limited to mainly cerebral white matter.^55^ [^18^F]-FPYBF-2 has also been used to study amyloid deposition in patients with atherosclerotic major cerebral artery disease.^56^

Flutafuranol ([^18^F]AZD4694 or [^18^F]-NAV4694) is a derivative of [^11^C]-PiB and targets β-amyloid fibrils. Consequently, the spatial distributions of [^11^C]-PiB and Flutafuranol are quite similar.^57^ Flutafuranol discriminates well between healthy controls and AD patients; AD patients typically show significantly increased binding in neocortical association areas and striatum compared to controls.^57,58^ Like many other amyloid tracers, Flutafuranol also shows nonspecific binding in white matter.^50,57^

Flortaucipir, also called T807 or [^18^F]AV-1451, binds to paired helical filament (PHF)-tau in neurofibrillary tangles (NFTs).^59,60^ It is currently the most widely used tau-PET tracer. Patients with AD show increased Flortaucipir binding relative to controls in the hippocampus and occipital, parietal, temporal, and frontal cortices, and binding increases with cognitive impairment and correlates with the severity of cortical atrophy.^61-63^ Flortaucipir uptake tends to be low in the cerebellum, making it a commonly used reference region.^60^ However, there are a number of identified sources of off-target binding, including neuromelanin and melanocytes in the choroid plexus, microhemorrhages, the substantia nigra, iron, and monoamine oxidase (MAO)-A.^64-67^ In addition to AD, Flortaucipir has been utilized in the study of other neurodegenerative diseases which have tau pathology, such as CTE, FTD, dementia with Lewy Bodies (DLB), and others.^64-66,68-70^

Florquinitau ([^18^F]-MK-6240) is a PET tracer that binds to tau aggregates.^71^ Its binding patterns are very similar to that of Flortaucipir, but it has been suggested to show greater range of SUVR values.^72^ Patients with AD show high uptake of Florquinitau relative to controls in the prefrontal cortex, posterior cingulate cortex, precuneus, inferior parietal cortex, lateral temporal cortex, and hippocampus.^71^ There is no difference in binding in the cerebellar gray matter, implicating the cerebellum as a useful reference region.^71^ Florquinitau does not exhibit nonspecific binding to MAO-A, unlike Flortaucipir, but does show nonspecific binding to the substantia nigra, melanocytes, and sites of hemorrhage.^73^

To date, a limited number of studies have been conducted using Aβ- and tau-PET to detect changes in AD-related protein deposition after acute or remote TBI in individuals at risk for AD. These studies are critical in characterizing the effects of TBI on AD biomarkers and risk for dementia in vivo. The purpose of this review is to synthesize the current body of literature relating to the use of Aβ and tau-PET in exploring the association between acute and remote TBI history and dementia pathological biomarkers in aging populations at risk for AD.

## Methods

### Eligibility criteria

Studies that used tau and/or Aβ-PET in human subjects to study the impact of remote or acute history of TBI in aging populations either with or at risk for AD were included. Gray literature (i.e., conference abstracts, meeting reports, etc.) were also eligible for inclusion. If a peer-reviewed and published paper was substantially similar to a gray literature report, the gray literature was excluded and the peer-reviewed paper was included. Exclusion criteria were: 1) concentration on a dementia other than AD, 2) no use of either Aβ or tau PET, 3) no specific focus on TBI, 4) studies using preclinical models, 5) study populations not old enough to be at risk for early- or late-onset AD (mean age <40 years), 6) studies not available in English, and 7) review or meta-analysis articles. Additionally, while many articles used different and/or interchangeable terminology to describe subjects’ injuries (e.g., TBI, head injury, concussion), for the purpose of this review, we will exclusively use the term TBI even if individual citations used different terminology.

### Information sources, search procedure, and study selection

A medical librarian composed and conducted comprehensive search strategies in MEDLINE (Ovid), Embase (Ovid), and the Cochrane Database of Systematic Reviews on January 9, 2023. Full search terms are available in the supplemental materials. The systematic review management tool Covidence removed duplicate records. 1476 unique citations were initially identified. The research team screened in two stages, first combined title and abstract screening and then full-text review. At each stage of the screening process, two reviewers independently screened each article and conflicts were resolved by consensus. After title and abstract screening, 81 articles were deemed appropriate for full-text review. After full-text review, the research team identified 24 articles that matched the inclusion criteria; data from these articles was then extracted using Covidence. Detailed data on the number of articles excluded during each step of the screening process can be found in Figure 1, and detailed data on the selected articles can be found in Table 1. No additional statistical analyses were performed on the selected citations, chiefly because of large variation in sample sizes across studies and the inability to identify subjects in commonly used datasets (e.g., the Alzheimer’s Disease Neuroimaging Initiative (ADNI)) which may have been included in the study sample of more than one citation.

**Fig. 1.**
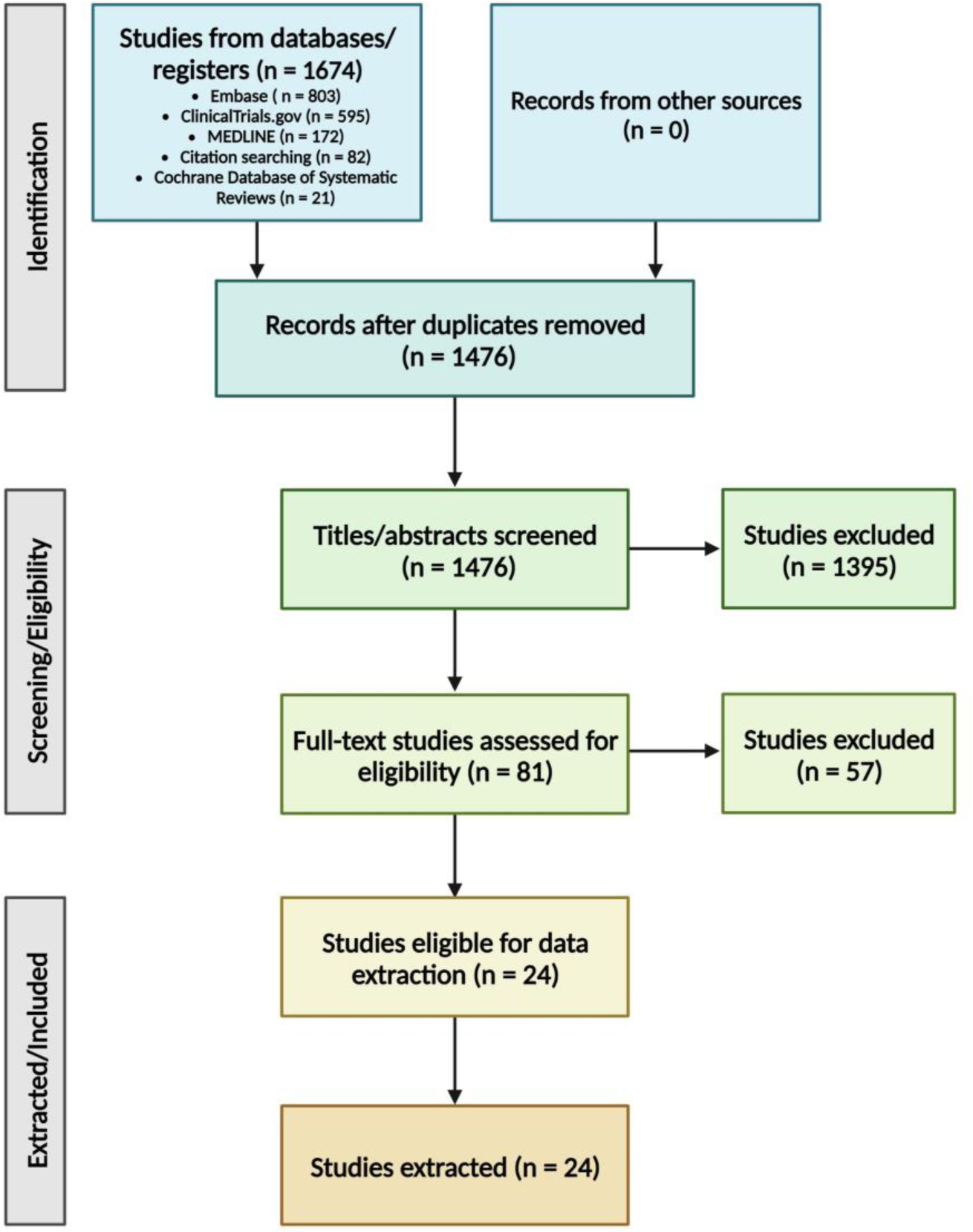
Preferred Reporting Items for Systematic Reviews and Meta-Analyses (PRISMA)^103^ flow diagram. Created with BioRender.com.

**Table 1.** Characteristics of included studies.

| Article | Total subjects | Pathology imaged | Tracer(s) | Subject sources | Injury/cognition breakdown | Cognitive statuses |
| --- | --- | --- | --- | --- | --- | --- |
| Asken 2021 | 134 | Amyloid | Florbetapir (92.5%), [<sup>11</sup>C]-PiB (7.5%) | UCSF Memory and Aging Center<br>Hillblom Aging Network | 48 mTBI (31 with 1 TBI, 17 with 2+), 86 non-TBI | CN |
| James 2021 | 499 | Amyloid | Florbetapir | Insight 46 (sub-study of National Survey of Health & Development, UK) | 80 with LOC HI > 15 years prior, 104 at least 1 HI at any time, 315 without HI | CN |
| Munro 2019 | 61 | Amyloid | Florbetapir | Dallas Lifespan Brain Study | - | CN |
| Wang 2017 | 90 | Amyloid | Florbetapir | ADNI | 45 with mTBI (8 normal, 10 preclinical AD, 17 MCI due to AD, 10 AD), 45 controls | CN and impaired |
| Weiner 2017 | 180 | Amyloid | Florbetapir | Vietnam Veterans identified by VA records (ADNI-DOD) | TBI only (22), PTSD only (63), both TBI + PTSD (32), and controls (63) | CN and impaired |
| Gatson 2016 | 2 | Amyloid | Florbetapir | Two clinic patients who had MVA | Both severe TBI and impaired | Case report |
| Mielke 2014 | 589 | Amyloid | [<sup>11</sup>C]-PiB | Mayo Clinic Study on Aging (MCSA) | 448 CN (74 TBI), 141 MCI (25 TBI) | CN and impaired |
| Mohamed 2018 | 164 | Amyloid | Florbetapir | ADNI-DOD Vietnam War veterans | 57 control, 21 TBI, 57 PTSD, 29 TBI + PTSD | CN |
| Mohamed 2022 | 241 | Amyloid | Florbetapir | ADNI | 19 TBI + CDR > 0.5, 22 TBI and CDR = 0, 100 without TBI and CDR > 0.5, 100 without TBI and CDR = 0 | CN and impaired |
| Schneider 2019 | 329 | Amyloid | Florbetapir | Atherosclerosis Risk in Communities-Positron Emission Tomography (ARIC-PET) Study | 81 with HI, 248 without HI | CN |
| Scott 2016 | 28 | Amyloid | [<sup>11</sup>C]-PiB | 9 clinic patients, recruited at least 11 months after their injury | 9 with TBI, 9 controls, 10 with AD | CN and impaired |
| Tateno 2015 | 1 | Amyloid | Florbetapir | Clinic patient struck by a vehicle | 1 with prior HI | Impaired |
| Ubukata 2020 | 70 | Amyloid | [<sup>18</sup>F]-FPYBF-2 | Recruited from outpatient clinic of neuropsychology unit at Dept. of Psychiatry & Department of Neurosurgery, Kyoto University Hospital | 20 chronic TBI patients, 50 controls | CN |
| Yang 2015 | 27 | Amyloid | Florbetapir | TBI patients treated for acute TBI at Taipei Medical University Hospital or TMU-Shuang Ho Hospital | 27 mTBI (6 with cognitive impairment/dementia), 10 controls | CN and impaired |
| Dore 2022 | 70 | Amyloid and tau | Florbetapir, Flortaucipir | Vietnam War veterans from Australian Imaging Biomarkers and Lifestyle Study of Aging-Veterans Study (AIBL-VETS) | 40 TBI, 30 controls | CN |
| Hicks 2022 | 146 | Both tau and amyloid | [<sup>18</sup>F]-NAV4694, [<sup>18</sup>F]-MK-6240 | Longitudinal head injury outcome study from admissions to inpatient rehabilitation program at Epworth HealthCare (Melbourne, AUS) | 87 TBI, 59 control | CN |
| Weiner 2022 | 289 | Amyloid and tau | Florbetapir, Flortaucipir | ADNI-DOD Vietnam War veterans | Baseline: 71 control, 81 PTSD, 43 TBI, 94 TBI + PTSD<br>Longitudinal: 60 control, 63 PTSD, 37 TBI, 65 TBI + PTSD | CN and impaired |
| Risacher 2021 | 752 | Amyloid and tau | Florbetapir, Florbetaben, Flortaucipir | ADNI and Indiana Memory and Aging Study (IMAS) | 302 impaired, 450 CN<br>63 with HI (38 CN, 25 impaired) | CN and impaired |
| Rowe 2019 | 128 | Tau | [<sup>18</sup>F]MK-6240 | - | 70 controls, 8 MCI, 10 AD, 4 other dementia, 36 remote TBI | CN and impaired |
| Gorgoraptis 2019 | 32 | Tau | Flortaucipir | 19 TBI from Institute of Health and Wellbeing, Head Injury Research Group, University of Glasgow; 2 TBI from clinic at Imperial College Healthcare NHS Trust | 21 TBI, 11 control | CN and impaired |
| Mohamed 2019 | 80 | Tau | Flortaucipir | ADNI-DOD Vietnam War veterans | 21 control, 10 TBI, 32 PTSD, 17 TBI + PTSD | CN and impaired |
| Mohamed 2022 | 120 | Tau | Flortaucipir | ADNI | 10 TBI + CDR > 0.5, 10 TBI and CDR = 0, 50 without TBI and CDR > 0.5, 50 without TBI and CDR = 0 | CN and impaired |
| Mountz 2017 | 13 | Tau | Flortaucipir | - | 7 chronic TBI, 6 control | CN |
| Mountz 2018 | 34 | Tau | Flortaucipir | - | 27 TBI (all impaired), 7 non-TBI controls | CN and impaired |

## Results and Discussion

### Amyloid

Of the 24 papers included in this review, 18 used at least one amyloid tracer. Florbetapir was used in 12 papers, [^11^C]-PiB was used in two papers, [^18^F]-FPYBF-2 was used in one paper, and [^18^F]-Flutafuranol was used in one paper. One paper used both Florbetapir and [^11^C]-PiB, and one paper used both Florbetapir and Florbetaben. The most commonly identified limitations were small sample size, limited clinical details about TBIs, the potential for recall bias, potentially biased recruitment strategies, and an inability to establish causation due to cross-sectional study designs.

### Evidence for limbic amyloid deposition

Four articles analyzed amyloid deposition in at least one limbic region (including the hippocampus, temporal pole, and entorhinal cortex) (Table 2)^74-77^.

**Table 2.** Amyloid deposition in limbic regions (- = not studied; NS = not significant, x = significant).

| **Article** | **Medial temporal lobe/hippocampus** | **Temporal pole** | **Entorhinal cortex** |
| --- | --- | --- | --- |
| Asken 2021 | - | - | - |
| Gatson 2016 | x | - | - |
| James 2021 | - | - | - |
| Mielke 2014 | - | - | - |
| Mohamed 2018 | - | - | - |
| Mohamed 2022 | - | x | - |
| Munro 2019 | - | - | - |
| Schneider 2019 | - | - | - |
| Scott 2016 | NS | - | - |
| Tateno 2015 | - | - | - |
| Ubukata 2020 | - | - | - |
| Wang 2017 | - | NS | NS |
| Weiner 2017 | - | - | - |
| Yang 2015 | - | - | - |
| Dore 2022 | - | - | - |
| Hicks 2022 | - | - | - |
| Weiner 2022 | - | - | - |
| Risacher 2021 | - | - | - |

### Hippocampus

Case 1 from Gatson and colleagues showed a decrease in amyloid in the bilateral hippocampus at the 12-month post-TBI scan compared to the 1-month scan.^74^ However, that patient then demonstrated a 15% increase in amyloid in the right hippocampus and no change in the left hippocampus at 24 months compared to 12 months.^74^ He also demonstrated worsened memory and cognitive outcomes at the 24-month time point compared to 12 months, suggesting his cognitive function may have been related to increased hippocampal amyloid deposition at that time point.^74^ This mixed outcome, where both amyloid clearance and accumulation are seen, suggests amyloid kinetics in the hippocampus after TBI may fluctuate over time after an injury. However, limited conclusions can be drawn, as this study reported only two cases. Case in point, Scott and colleagues (2016) observed no significant difference in hippocampal [^11^C]-PiB binding in a sample of nine subjects with TBI.^76^ Together, these two studies provide disparate evidence regarding amyloid deposition in the hippocampus after TBI. However, as only 11 subjects total were examined, replications are needed to further explore how TBI may be associated with hippocampal amyloid deposition.

### Temporal pole/entorhinal cortex

Mohamed and colleagues (2022) found evidence for increased amyloid in the temporal pole of Aβ-positive (Aβ+) subjects with TBI compared to those without TBI, but no difference in cognition.^75^ This suggests that if there is an association of TBI with amyloid deposition in the temporal pole, there may not be a consequential cognitive impact. Contrastingly, Wang and colleagues (2017) found no significant difference in Florbetapir binding in the temporal pole or entorhinal cortex between subjects with and without TBI.^77^ Limited sample size may have contributed to these mixed findings, as both studies used a relatively small number of subjects with TBI. It is also plausible that exclusive examination of only mild TBI subjects contributed to the lack of conclusive findings regarding amyloid deposition in the temporal pole. The similar lack of support for a cognitive impact of amyloid deposition in this region may also be attributable to examination of mild TBI only. Future studies utilizing a greater number of subjects, as well as subjects with more severe injuries, will be necessary to confirm whether TBI may associate with deposition of amyloid in limbic brain areas, and if this can impact cognition.

### Evidence for subcortical amyloid deposition

Six articles investigated amyloid deposition in at least one subcortical region of interest (including the thalamus, globus pallidus, caudate and putamen (striatum), and cerebellum); global white matter analysis was also included in this category (Table 3).^74-76,78-80^

**Table 3.** Amyloid deposition in subcortical regions.

| **Article** | **Thalamus** | **Globus pallidus** | **Caudate** | **Putamen** | **Cerebellum** | **White matter** |
| --- | --- | --- | --- | --- | --- | --- |
| Asken 2021 | - | - | - | - | - | - |
| Gatson 2016 | NS | - | x | x | - | - |
| James 2021 | - | - | - | - | - | - |
| Mielke 2014 | - | - | - | - | - | - |
| Mohamed 2018 | - | - | - | - | x | - |
| Mohamed 2022 | - | - | - | - | x | - |
| Munro 2019 | - | - | - | - | - | - |
| Schneider 2019 | - | - | - | - | - | - |
| Scott 2016 | NS | - | NS | NS | x | - |
| Tateno 2015 | - | - | - | - | - | - |
| Ubukata 2020 | NS | NS | NS | NS | - | x |
| Wang 2017 | - | - | - | - | - | - |
| Weiner 2017 | - | - | - | - | - | - |
| Yang 2015 | - | - | x | x | NS | - |
| Dore 2022 | - | - | - | - | - | - |
| Hicks 2022 | - | - | - | - | - | - |
| Weiner 2022 | - | - | - | - | - | - |
| Risacher 2021 | - | - | - | - | - | - |

### Striatum (caudate and putamen)

Case 1 from Gatson and colleagues demonstrated decreased amyloid in the caudate at the 12-month Florbetapir scan compared to the 1-month time point, and no change at 24 months.^74^ However, Case 2 showed increased amyloid in the left anterior putamen and decreased amyloid in the left caudate at 12 months compared to 1 month.^74^ Case 2 then had decreased amyloid at 24 months compared to 12 months in the left anterior putamen.^74^ The results from Case 2 suggest a possible rise in striatal amyloid deposition shortly after TBI, followed by eventual clearance; however, this patient was relatively young (37 at the time of TBI), so these results may not be generalizable to significantly older populations. Nonetheless, these findings, along with previously described findings from limbic regions, provide further evidence that amyloid deposition patterns after TBI are dynamic in the relatively acute period after injury. Intriguingly, neither patient demonstrated a change in the level of binding in the thalamus at any time point, which may indicate the thalamus is relatively spared from TBI-associated amyloid deposition. However, this data should be interpreted with caution, as only two subjects were studied.

Yang and colleagues (2015) found mTBI subjects with dementia (mTBI + D) showed high Florbetapir uptake in the bilateral striatum relative to mTBI subjects without dementia (mTBI – D) and healthy control subjects.^80^ However, *APOE* ε4 allele frequency was higher in the mTBI + D group compared to controls, which may have resulted in biased comparisons, as *APOE* ε4 is associated with elevated amyloid burden.^81^ Additionally, though the findings point to increased striatal amyloid in the mTBI + D group, this may be a consequence of the presence of AD rather than TBI, as the mTBI – D group showed no significant difference in striatal amyloid compared to healthy controls. Furthermore, Ubukata and colleagues (2020) found patients with chronic diffuse axonal injury (DAI), a severe form of TBI associated with widespread axonal damage and the potential for coma or death,^82^ showed non-significantly increased binding in subcortical regions compared to controls, but patients with chronic focal injuries showed lower binding in all subcortical ROIs compared to DAI patients and no change in binding compared to controls.^79^ This suggests the striatum may not be vulnerable to amyloid deposition after TBI, except in extreme cases of TBI where DAI is present. Together, the results indicate striatal amyloid deposition may be more closely linked to dementia or severe TBI than mild TBI. However, as with other previously described brain regions, future studies in expanded samples with a range of TBI severities will be necessary to clarify these findings.

### Cerebellum

Two studies from Mohamed and colleagues found evidence for increased amyloid deposition in the cerebellum in patients with TBI.^75,78^ The group published results (Mohamed and colleagues, 2018) suggesting veterans with TBI had higher Florbetapir uptake compared to controls in the cerebellar tonsil.^78^ They noted in a subsequent report (Mohamed and colleagues, 2022) that Aβ-positive (Aβ+) subjects with history of TBI also had higher Florbetapir retention in the cerebellum compared to individuals without history of TBI.^75^ Importantly, these findings suggest the use of the cerebellum as a reference region in PET studies may be problematic if the cerebellum is vulnerable to amyloid deposition after TBI. However, it is worth noting that a significant corrigendum to their 2018 study was published in response to a letter to the editor expressing concerns over a failed attempt to replicate the study.^83,84^ Even after the corrigendum, the primary conclusions of the study were still unable to be replicated, so these results may or may not accurately reflect group-wise differences in this population.^84^ However, in support of these findings, Scott and colleagues also observed that patients with TBI showed increased cerebellar [^11^C]-PiB binding, adding evidence that the cerebellum may not be an ideal reference region for amyloid-PET studies in TBI. Intriguingly, the patients with AD in that study did not show the same increased binding in the cerebellum that TBI patients did. This suggests TBI may be associated with a unique cerebellar amyloid deposition pattern that is not seen in typical AD.^76^ In contrast to these results, Yang and colleagues found no evidence for elevated cerebellar Florbetapir binding in 27 mTBI patients compared to controls.^80^ Nevertheless, the finding from three studies of increased amyloid tracer binding in the cerebellum is notable. This demands further exploration and replication in larger datasets to determine whether the evidence for elevated amyloid tracer uptake in the cerebellum of TBI subjects truly reflects TBI-associated amyloid deposition, as this finding may influence future selection of reference regions in subsequent studies investigating amyloid deposition in subjects with known history of TBI.

### White matter

Ubukata and colleagues (2020) found chronic TBI patients with DAI had higher [^18^F]- FPYBF-2 binding in white matter compared to controls, but saw no difference between focal TBI patients and controls.^79^ These results suggest if TBI is a risk factor for amyloid deposition in the white matter, it is likely limited to more severe injury cases that include axonal injury. However, limited conclusions can be drawn without replication, as this was the only study to examine amyloid deposition in global white matter after TBI.

### Evidence for cortical amyloid deposition

All 18 articles which utilized an amyloid tracer assessed at least one measure of cortical amyloid deposition. These ROIs included measures of global amyloid status and/or burden, as well as the frontal, parietal, temporal, and occipital lobes, the corpus callosum and/or fornix, the anterior and the posterior cingulate, and the cuneus or precuneus (Table 4).^74-80,85-95^

**Table 4.** Amyloid deposition in cortical regions.

| **Article** | **Global amyloid status/burden** | **Frontal lobe** | **Parietal lobe** | **Temporal lobe** | **Occipital lobe** | **Corpus callosum** | **Ant. cingulate** | **Post. cingulate** | **Pre/cuneus** |
| --- | --- | --- | --- | --- | --- | --- | --- | --- | --- |
| Asken 2021 | NS | - | - | - | - | - | - | - | - |
| Gatson 2016 | - | NS | NS | NS | x | - | NS | NS | x |
| James 2021 | NS | - | - | - | - | - | - | - | - |
| Mielke 2014 | x | - | - | - | - | - | - | - | - |
| Mohamed 2018 | - | x | x | x | - | x | x | x | x |
| Mohamed 2022 | - | x | NS | x | - | - | x | x | x |
| Munro 2019 | NS | - | - | - | - | - | - | - | - |
| Schneider 2019 | x | x | NS | NS | NS | - | NS | x | NS |
| Scott 2016 | - | NS | - | - | NS | - | NS | x | x |
| Tateno 2015 | x | - | - | - | - | - | - | - | - |
| Ubukata 2020 | - | NS | NS | x | x | NS | - | - | - |
| Wang 2017 | - | - | NS | NS | - | - | - | NS | NS |
| Weiner 2017 | NS | - | - | - | - | - | - | - | - |
| Yang 2015 | x | x | x | x | NS | - | - | NS | x |
| Dore 2022 | NS | - | - | - | - | - | - | - | - |
| Hicks 2022 | NS | - | - | - | - | - | - | - | - |
| Weiner 2022 | NS | - | - | - | - | - | - | - | - |
| Risacher 2021 | NS | - | - | - | - | - | - | - | - |

### Global amyloid status and/or burden

Schneider and colleagues (2019) found history of TBI to be associated with increased global cortical Florbetapir SUVRs in cognitively normal individuals.^89^ Mielke and colleagues (2014) similarly observed no association between TBI and amyloid deposition in cognitively normal individuals via [^11^C]-PiB, but amongst individuals with mild cognitive impairment (MCI), those with TBI had ∼18% higher global amyloid and 5-fold higher odds of increased amyloid compared to those without TBI.^87^ Additionally, Yang and colleagues found increased amyloid in mTBI + D patients, but no difference in between mTBI - D and controls.^80^ Together, these studies provide evidence for an association of TBI with cortical amyloid deposition, but suggest it may be predominantly driven by cognitively impaired individuals. In line with this, Tateno et al. (2015) published a case report describing the clinical progression of a 48-year-old woman (APOE ε4/ε4) who was hit by a car at age 43.^91^ She presented to the clinic with new onset of memory symptoms that worsened rapidly, had an AD-like pattern of amyloid accumulation on Florbetapir-PET, and was subsequently diagnosed with MCI which progressed to AD three years after the accident.^91^ Though causation cannot be determined, the appearance of MCI shortly after TBI and the patient’s eventual diagnosis of AD indicate that the onset of cognitive impairment might have been related to the TBI. It is plausible that the patient’s injury induced a rise in amyloid deposition, which had likely already been occurring for some time, and this triggered her rapid cognitive deterioration.

Contrastingly, Asken and colleagues (2021) found no association between mTBI and cortical Aβ burden or likelihood of being positive on Florbetapir or [^11^C]-PiB -PET, and no interaction between mTBI and Aβ burden on cognition. Longitudinal data from a subset of subjects with a second Aβ PET scan suggested remote mTBI might be associated with faster cortical Aβ burden increase over time, but this was not statistically significant.^85^ James and colleagues (2021) also observed no associations between TBI with loss of consciousness (LOC) and amyloid status; however, TBI with LOC was associated with lower cognitive function on the digit symbol substitution test (DSST).^86^ Evidence of a link between TBI with LOC and cognitive changes, but no association with amyloid deposition on PET, is somewhat contrary to the study by Asken and colleagues (2021) which found no evidence of an impact of TBI on either cognition or amyloid.^86^ However, that study examined only mTBI subjects, which suggests LOC may be a better predictor of dementia-related cognitive changes than simply the presence or absence of a TBI. LOC status can be used as a proxy for injury severity, and injuries resulting in LOC may be considered more severe. The study from James and colleagues included 80 subjects with LOC, likely resulting in their sample including more severe injuries. This may explain their finding of a link between TBI with LOC and cognitive changes, and also supports the notion that increased injury severity correlates with increased amyloid deposition, which would be expected to track with cognitive decline.

Similar null results were reported in Munro et al. (2019), where a linear regression model assessing whether mTBI could predict the rate of amyloid accumulation was significant, but prior mTBI was not independently predictive of amyloid accumulation.^88^ These results were reported as part of a conference abstract submitted for the 2019 Alzheimer’s Association International Conference, so they have not been subjected to extensive peer review, and more detailed information about subjects’ characteristics and study methodologies is not available. However, the findings are shared by other studies, including Hicks et al. (2022) which also saw no association of TBI with Aβ burden.^93^ Furthermore, Risacher et al. (2021) found no significant effect of TBI on amyloid, though there was an interaction between amyloid positivity and TBI with LOC where Aβ+ subjects with TBI with LOC had the highest tau deposition. This adds further evidence supporting the notion that severe TBI may be more likely to result in increased AD-related pathology than mild TBI.^95^

A select number of studies examined the effects of TBI on global amyloid deposition in former military servicemembers. Weiner et al. (2017) found no evidence for an association between TBI and increased brain amyloid in Vietnam War veterans.^90^ A group of subjects with both TBI and a diagnosis of post-traumatic stress disorder (PTSD) (TBI + PTSD group) showed slightly lower superior parietal volumes and lower scores on the Clinical Dementia Rating Scale Sum of Boxes (CDR-SB) relative to controls, but there was no association between TBI and elevated amyloid deposition.^90^ These results suggest cognitive decline in some veterans with history of TBI may be a consequence of concurrent PTSD rather than TBI alone, since this result was only observed in the TBI + PTSD group and not the TBI-only group. A 2022 study from the same group found similar results, where exposure groups (TBI, PTSD, and TBI + PTSD) had slightly higher MCI incidence, but no significant differences in Florbetapir SUVR.^94^

In line with those findings, Dore et al. (2022) also found no difference in Florbetaben uptake between TBI and non-TBI veteran subjects.^92^ There were still no significant differences in amyloid levels when the TBI group was subdivided into mTBI (n=15) and moderate-to-severe TBI (n=25).^92^ As this study only examined cognitively normal subjects, the validity of the previously-observed association between TBI + PTSD and worsened cognitive performance cannot be assessed, but the lack of evidence for increased amyloid in TBI patients is congruent with both studies from Weiner and colleagues.

Altogether, these 12 studies provide mixed results regarding an association of TBI with cortical amyloid deposition. However, it is plausible that regional changes in amyloid differences were unable to be detected in reports that only examined a summary cortical ROI. From the citations which do support an association between TBI and cortical amyloid deposition, there is evidence to suggest this may be driven by individuals who had either a more severe TBI (i.e., experienced LOC) or ongoing cognitive impairment. These possibilities provide an avenue for future research to both replicate and better characterize these observations.

### Frontal lobe

Schneider et al. (2019) found an association of TBI with elevated amyloid in the orbitofrontal cortex (OFC), prefrontal cortex (PFC), and superior frontal cortex (SFC).^89^ Similarly, Yang et al. (2015) showed subjects with mTBI and ongoing cognitive impairment (mTBI + D) had higher Florbetapir uptake in frontal regions relative to controls.^80^ Additionally, in the study from Schneider and colleagues, subjects with more than one TBI demonstrated elevated Florbetapir SUVRs in the PFC and SFC compared to subjects with only one TBI.^89^ This indicates there may be a dose-dependent effect of TBI on amyloid levels in the frontal lobe, whereby having more than one TBI results in elevated amyloid compared to a single injury.

Intriguingly, Mohamed and colleagues (2018) showed TBI subjects had higher Florbetapir uptake compared to controls in the supplementary motor area (SMA), but lower uptake in ventrolateral PFC.^78^ The TBI + PTSD group also demonstrated lower uptake in the SFC.^78^ Their 2022 study then showed Aβ+ subjects with history of TBI demonstrated higher Aβ deposition in the medial frontal cortex (dorsal and ventral), OFC, inferior frontal gyrus, and right SFC.^75^ These results are somewhat contrasting, finding evidence for elevated amyloid in some frontal regions and lower amyloid in others. To add further complexity, Scott and colleagues (2016) reported that patients with TBI did not show significant differences in frontal lobe amyloid compared to controls, but there was a group-by-region interaction driven partly by decreased [^11^C]-PiB binding in the SFC.^76^ Additionally, Ubukata and colleagues (2020) found chronic DAI patients showed non-significantly increased binding in frontal regions compared to controls, but focal injury patients showed lower [^18^F]-FPYBF-2 binding in all frontal ROIs compared to DAI patients and no change relative to controls.^79^ These mixed results indicate the effect of TBI on frontal lobe amyloid deposition is heterogeneous, and may differ depending on the particular frontal lobe ROI being examined. Furthermore, there may be differential effects of injury severity, as well as the number of injuries sustained. Future studies should individually investigate various sub-ROIs within the frontal lobe with greater detail to clarify these findings.

### Parietal lobe

The TBI + PTSD group from Mohamed and colleagues (2018) demonstrated low Florbetapir uptake in the inferior parietal cortex.^78^ Contrastingly, however, the same group showed in a 2022 study that Aβ+ subjects with TBI had higher amyloid deposition compared to individuals without TBI in the supramarginal gyrus.^75^ Yang et al. (2015) also found mTBI + D subjects showed higher Florbetapir uptake in the right parietal regions relative to controls and the mTBI – D group,^80^ and Wang and colleagues (2017) demonstrated mTBI history was associated with decreased cortical thickness in the superior parietal cortices in subjects with preclinical AD.^77^ Finally, Ubukata and colleagues (2020) noted that DAI patients showed non-significantly increased [^18^F]-FPYBF-2 binding in parietal regions compared to controls,^79^ but focal injury patients showed lower parietal ROI binding compared to DAI patients and no change relative to controls.^79^ Findings from these studies are somewhat varied, but point to a general trend of evidence for altered parietal lobe amyloid deposition after TBI, with the potential for varying effects based on injury severity and the presence of psychiatric comorbidities like PTSD.

### Temporal lobe

The TBI + PTSD group studied by Mohamed et al. (2018) showed higher Florbetapir uptake relative to controls in the left medial temporal gyrus, and lower uptake in the superior temporal gyrus (STG).^78^ However, their 2022 study indicated Aβ+ subjects with TBI had higher Aβ deposition compared to individuals without TBI in the posterior STG.^75^ As previously discussed, this is further evidence that a comorbid PTSD diagnosis in conjunction with TBI history may be associated with altered amyloid levels. Yang et al. (2015) also found mTBI + D subjects showed high Florbetapir uptake in the bilateral STG relative to controls.^80^ Furthermore, Ubukata and colleagues (2020) observed that patients with DAI showed significantly increased binding in the temporal cortices relative to both controls and focal injury patients, but focal injury patients showed no change compared to controls.^79^ This reinforces the idea that increased amyloid deposition may be more likely in severe TBI than mTBI. Contrastingly, Wang and colleagues (2017) found no significant differences in Florbetapir SUVR in the temporal lobes between the mTBI group and non-TBI group.^96^ Once again, mixed results point to the need for future studies in expanded samples to clarify the previously published findings, and to investigate the differential effects of varying injury severities and comorbid psychiatric conditions on amyloid deposition in the temporal lobe.

### Occipital lobe

There are very incongruent results regarding amyloid deposition in the occipital lobe after TBI. At both 12 and 24 months post-injury, Case 2 from Gatson and colleagues had lower amyloid in the left occipital cortex relative to the previous scan timepoint.^74^ However, Scott and colleagues (2016) found no difference in amyloid deposition in the occipital lobes in patients with TBI compared to controls.^76^ Contrastingly, Mohamed and colleagues (2022) reported Aβ+ subjects with TBI did in fact have higher Aβ deposition compared to those without TBI in the lingual and fusiform gyri,^75^ and Ubukata and colleagues (2020) found chronic DAI subjects had increased [^18^F]-FPYBF-2 binding in the occipital cortices compared to controls.^79^ Every potential trend in amyloid deposition after TBI in the occipital cortex was observed: one study found no evidence for altered amyloid levels, two studies observed increased amyloid, and one case report noted a decrease in amyloid. These disparate findings suggest the patterns of amyloid deposition in the occipital cortex may be highly subject-specific, and could be related to TBI severity. Additionally, small sample sizes and use of different occipital ROIs may also have contributed to the contrasting findings. As with many previously discussed regions, replications are necessary.

### Corpus callosum

The TBI + PTSD group from Mohamed and colleagues (2018) demonstrated increased Florbetapir uptake relative to controls in the corpus callosum.^78^ Contrastingly, Ubukata and colleagues (2020) found no significant differences in [^18^F]-FPYBF-2 binding in the corpus callosum.^79^ These data should be interpreted with extreme caution, as off-target binding of amyloid tracers to white matter must be considered as a potential source of false positives.^42,43^ Future replications are needed to clarify these findings and better characterize off-target binding of amyloid-PET tracers in white matter tracts.

### Cingulate (anterior and posterior)

Schneider et al. (2019) found evidence for elevated amyloid in the posterior cingulate cortex (PCC) in TBI subjects.^89^ Contrastingly, Wang and colleagues (2017) saw no difference in mean Florbetapir SUVR between mTBI and non-TBI subjects in the PCC. However, in line with Schneider and colleagues, the TBI + PTSD group from Mohamed and colleagues (2018) also demonstrated increased Florbetapir uptake relative to controls in the cingulate cortex, and the 2022 study from that group reported Aβ+ subjects with TBI had higher amyloid deposition compared to individuals TBI in both the anterior cingulate cortex (ACC) and PCC.^75^ Intriguingly, Scott and colleagues (2016) found that patients with TBI as well as those with AD exhibited increased amyloid compared to controls in the PCC, but not the ACC.^76^ Furthermore, Yang et al. (2015) reported higher Florbetapir uptake in the mTBI + D group relative to controls in the right cingulate.^80^ These results generally point to both the ACC and PCC as being vulnerable to amyloid deposition in subjects with TBI. However, as with other brain regions, repetitions of these studies and considerations of varying injury severity are imperative.

### Cuneus and precuneus

In case 2 from Gatson and colleagues, there was decreased amyloid in the precuneus at both 12 and 24 months compared to the previous scan timepoint.^74^ However, Mohamed and colleagues (2018) reported that TBI subjects had higher Florbetapir uptake compared to controls in the precuneus.^78^ Additionally, that group showed in 2022 that Aβ+ subjects with TBI had higher Aβ deposition compared to those without TBI in the precuneus and cuneus.^75^ Scott and colleagues (2016) also published evidence for increased precuneus amyloid levels in patients with TBI and patients with AD compared to controls.^76^ In addition, the mTBI +D group from Yang et al. (2015) exhibited high Florbetapir uptake in the left cuneus relative to controls, and higher uptake in the bilateral precuneus compared to the mTBI – D group.^80^ There was also increased amyloid in the right cuneus between controls and mTBI - D subjects, and higher amyloid in the bilateral precuneus in mTBI + D relative to mTBI – D.^80^ The difference between the mTBI + D and mTBI – D groups is likely attributable to dementia status, but this study also provides evidence for greater cuneus amyloid burden in the mTBI – D group relative to controls, which suggests an effect of TBI alone.^80^ Furthermore, Wang and colleagues (2017) reported that mTBI history was associated with decreased precuneus thickness in preclinical AD subjects.^77^

Taken together, the majority of these results indicate the cuneus/precuneus may be susceptible to amyloid deposition after TBI. Though both cases discussed by Gatson and colleagues showed decreased amyloid in the precuneus, there are a number of potential explanations for this. First, the TBIs experienced by those cases were more recent injuries than those described in other studies, as this study focused on the relatively acute time period (24 months) after severe TBI. Additionally, one of the cases was comparatively young, and might therefore demonstrate different amyloid kinetics than older subjects. It is thus possible that the contrasting findings reported by Gatson and colleagues are simply a reflection of methodological differences. The remaining reports present relatively clear evidence in support of increased amyloid in the cuneus/precuneus in subjects with TBI. However, replication of these studies that take into consideration both age at injury and time since injury should be conducted, and will be informative in clarifying these observations.

### Tau

Of the 24 papers discussed in this review, 10 used a tau tracer. Eight papers used Flortaucipir, and two papers used [^18^F]-MK-6240. The major limitations identified in the citations were small numbers of participants with TBI history, a lack of objective clinical details, and the potential for recall bias.

### Evidence for limbic tau deposition

Six papers examined patterns of tau deposition in at least one limbic ROI, including the medial temporal lobe/hippocampus, entorhinal cortex, and temporal pole (Table 5).^94,95,97-100^

**Table 5.** Tau deposition in limbic regions.

| **Article** | **Medial temporal lobe/hippocampus** | **Temporal pole** | **Entorhinal cortex** |
| --- | --- | --- | --- |
| Dore 2022 | - | - | - |
| Hicks 2022 | - | - | - |
| Weiner 2022 | - | - | NS |
| Risacher 2021 | x | - | - |
| Gorgoraptis 2019 | NS | - | - |
| Mohamed 2019 | - | x | - |
| Mohamed 2022 | x | x | x |
| Mountz 2017 | x | - | - |
| Mountz 2018 | - | - | - |
| Rowe 2019 | - | - | - |

### Medial temporal lobe/hippocampus

Gorgoraptis and colleagues (2019) saw no significant difference in Flortaucipir binding in a medial temporal composite ROI incorporating the entorhinal, perirhinal, and parahippocampal cortices, and hippocampus.^97^ However, composite ROIs may conceal region-specific effects that could be identified by examination of individual ROIs. In support of this possibility, Risacher and colleagues (2021) found evidence for increased tau in the medial temporal lobe (MTL) of adults with TBI with or without LOC.^95^ Furthermore, in Mohamed and colleagues (2022), both cognitively normal and impaired subjects with TBI exhibited increased Flortaucipir SUVR in the hippocampus compared to those without TBI.^98^ Additionally, TBI subject 3 from Mountz and colleagues (2017) demonstrated diffusely increased Flortaucipir uptake in the hippocampus.^100^ This data is from a conference abstract that has not been subjected to extensive peer review, and should be interpreted with caution. Nevertheless, these reports together present relatively consistent evidence for elevated tau in the hippocampus/medial temporal lobe in subjects with TBI.

### Entorhinal cortex

Weiner and colleagues (2022) reported slightly higher incidences of MCI in exposure groups (TBI, PTSD, and TBI + PTSD), but no significant differences in entorhinal cortex Flortaucipir SUVRs either cross-sectionally or longitudinally.^94^ Oppositely, Mohamed and colleagues (2022) found that cognitively normal subjects with TBI had elevated tau in the transentorhinal cortex compared to those without TBI.^98^ No clear consensus can be determined from these two studies, and replications are necessary. Future studies should carefully consider TBI history and tau levels in the entorhinal cortex among both cognitively normal and impaired subjects, as previously described results have shown the effect of TBI on AD-related protein deposition may be driven by impaired subjects.

### Temporal pole

In Mohamed and colleagues (2019), subjects with TBI showed higher mean Flortaucipir SUVR in the temporal pole compared to control subjects without TBI.^99^ In the 2022 paper from the same group, cognitively normal subjects with TBI showed increased tau in the temporal pole compared to those without TBI.^98^ This is intriguing initial evidence for a link between TBI history and temporal pole tau deposition, but replication by other groups and within other datasets is necessary to strengthen the observations.

### Evidence for cortical and/or subcortical tau deposition

All ten papers analyzed at least one cortical and/or subcortical ROI, including cortical composite ROIs (mesial-temporal, temporoparietal, meta-temporal, and rest of neocortex composites), frontal, temporal, parietal, and occipital lobes, the cingulate gyrus, precuneus, insula, basal ganglia, and substantia nigra (Table 6).^92-95,97-102^

**Table 6.** Tau deposition in cortical and sub-cortical regions.

| Article | Mesial-temporal composite<br>(entorhinal cortex, hippocampus, parahippocampus, amygdala) | Temporoparietal composite<br>(inferior and middle temporal, fusiform, supramarginal and angular gyri, posterior cingulate/precuneus, superior and inferior parietal, lateral occipital) | Meta-temporal composite<br>(entorhinal cortex, hippocampus proper, parahippocampus, amygdala, fusiform, inferior and middle temporal gyri, temporo-occipital region, angular gyrus) | Rest of neocortex composite<br>(dorsolateral and ventrolateral prefrontal, orbitofrontal, gyrus rectus, superior temporal and anterior cingulate) | Frontal lobe | Temporal lobe | Parietal lobe | Occipital lobe | Cingulate gyrus | Precuneus | Insula | Basal ganglia | Substantia nigra |
| --- | --- | --- | --- | --- | --- | --- | --- | --- | --- | --- | --- | --- | --- |
| Dore 2022 | NS | NS | - | NS | - | - | - | - | - | - | - | - | - |
| Hicks 2022 | x | x | x | x | - | - | - | - | - | - | - | - | - |
| Weiner 2022 | - | - | - | - | - | NS | - | - | - | - | - | - | - |
| Risacher 2021 | - | - | - | - | x | x | x | - | - | x | - | - | - |
| Gorgoraptis 2019 | - | - | - | - | - | - | - | x | - | - | - | - | - |
| Mohamed 2019 | - | - | - | - | x | x | x | NS | - | x | x | x | - |
| Mohamed 2022 | - | - | - | - | - | x | x | x | - | x | - | - | - |
| Mountz 2017 | - | - | - | - | x | x | x | x | - | x | - | x | x |
| Mountz 2018 | - | - | - | - | - | - | x | x | x | x | - | - | - |
| Rowe 2019 | x | x | - | - | - | - | - | - | - | - | - | - | - |

### Cortical composite ROIs

Dore and colleagues (2022) saw no difference in Flortaucipir uptake between TBI and non-TBI subjects in any of three composite ROIs (mesial-temporal, temporoparietal, and rest of neocortex).^92^ There were also no differences upon stratifying by injury severity (mTBI versus mild-to-moderate TBI).^92^ Using the same three composite ROIs with the addition of a meta-temporal composite ROI, Hicks and colleagues (2022) similarly noted no association of TBI with elevated [^18^F]-MK-6240 SUVR; intriguingly, control subjects actually demonstrated higher [^18^F]-MK-6240 SUVRs.^93^ Furthermore, Rowe and colleagues (2019) observed no evidence for increased tau pathology in similar temporoparietal and mesial-temporal composite ROIs in subjects with TBI compared to Aβ+ patients with AD, Aβ+ patients with MCI, or Aβ+ cognitively normal patients.^102^ However, this is data from a meeting report that has not been subjected to extensive peer-review, so should be interpreted with caution. Nonetheless, negative results from all studies suggest no apparent association between TBI and amyloid deposition in cortical composite ROIs. However, as mentioned previously, composite ROIs may conceal regional alterations to tau deposition patterns in subjects with TBI. Therefore, these observations must be considered alongside studies which examined individual cortical ROIs, findings from which will be subsequently discussed.

### Frontal lobe

Risacher and colleagues (2021) observed increased tau in the frontal lobe of adults with TBI with or without LOC.^95^ However, these findings were driven by individuals with cognitive impairment and there were no effects in cognitively normal subjects.^95^ Similarly, Mohamed and colleagues (2019) found subjects with TBI showed significantly higher mean Flortaucipir SUVR in the superior frontal gyrus, middle frontal gyrus, medial OFC, lateral OFC, and precentral gyrus compared to control subjects without TBI.^99^ The TBI + PTSD group also showed higher mean Flortaucipir SUVR in the middle frontal gyrus compared to control subjects without TBI, again suggesting a possible interaction of psychiatric comorbidities on frontal lobe tau deposition in subjects with TBI.^99^ Furthermore, TBI subject 2 from Mountz and colleagues (2017) had intense Flortaucipir uptake in the bilateral OFC, and TBI subject 3 had diffusely increased Flortaucipir uptake in the OFC.^100^ These results offer corresponding evidence that tau may be elevated in frontal lobe ROIs in subjects with TBI. Furthermore, these studies suggest the association of TBI with frontal lobe tau deposition may be stronger in individuals with cognitive impairment and/or psychiatric comorbidities. Future studies should seek to further define the differential impacts of cognitive impairment and psychiatric comorbidities such as PTSD on tau deposition patterns in frontal lobe gyri of subjects with TBI.

### Temporal lobe

Weiner and colleagues (2022) saw no evidence for increased tau in a temporal lobe composite ROI over cross-sectional or longitudinal analyses using Flortaucipir.^94^ However, TBI subjects from Mohamed and colleagues (2019) showed significantly higher mean Flortaucipir SUVRs in the transverse temporal gyrus, but lower SUVRs in the left inferior temporal gyrus, compared to control subjects without TBI.^99^ The TBI + PTSD group also exhibited higher mean Flortaucipir SUVR in the transverse and superior temporal gyri compared to control subjects without TBI.^99^ Furthermore, Mohamed and colleagues (2022) reported cognitively normal subjects with TBI had elevated tau in the inferior temporal gyrus, middle temporal gyrus, and superior temporal gyrus compared to those without TBI.^98^ Additionally, cognitively impaired subjects with TBI demonstrated increased SUVR in the inferior temporal gyrus and middle temporal gyrus compared to those with no history of TBI.^98^ To add to this evidence, TBI subject 2 from Mountz and colleagues (2017) showed higher Flortaucipir uptake in the left medial temporal cortex, and TBI subject 3 had diffusely increased uptake in the lateral temporal lobe.^100^

Together, these studies generally support an association between TBI and temporal lobe tau deposition. Additionally, the results also support the observation that subjects with concurrent cognitive impairment and/or PTSD may exhibit differential effects of TBI on protein deposition. While the studies that used individual gyrus ROIs did note associations between TBI and temporal tau deposition, Weiner and colleagues may not have identified significant differences due to the use of a temporal composite ROI that may have concealed gyrus-specific effects. As discussed previously, future studies should carefully consider the tradeoffs of composite ROIs in studying regional changes in protein deposition in subjects with TBI.

### Parietal lobe

Risacher and colleagues (2021) reported greater tau in the parietal lobe of adults with history of TBI independent of LOC status.^95^ Mohamed and colleagues (2019) also showed significantly higher mean Flortaucipir SUVR in the postcentral gyrus and supramarginal gyrus in TBI subjects compared to control subjects without TBI.^99^ Interestingly, the TBI + PTSD group also exhibited higher mean SUVR in the supramarginal gyrus compared to control subjects without TBI.^99^ In their 2022 study, the same group showed cognitively normal subjects with TBI had increased tau in the angular gyrus, inferior parietal gyrus, and superior parietal gyrus compared to those who had no history of TBI.^98^ Furthermore, cognitively impaired subjects with TBI showed increased SUVR in the inferior parietal gyrus compared to those with no history of TBI.^98^ Additionally, TBI subject 3 from Mountz and colleagues (2017) had elevated Flortaucipir in the bilateral parietal lobe,^100^ and in a report of 27 TBI subjects and 7 cognitively normal controls, Mountz and colleagues (2018) observed that the three TBI patients who had numerous (greater than 10) injuries had high Flortaucipir uptake in several cortical grey matter regions, including the parietal lobes.^101^ However, this data was reported in a meeting report, and has not been subjected to peer review, and thus should be interpreted with caution. In spite of this, these five studies agree that there is evidence for elevated tau in a number of parietal lobe ROIs in subjects who have experienced a TBI. Replications in each of these ROIs will be necessary to add strength to the findings.

### Occipital lobe

Gorgoraptis and colleagues (2019) observed elevated Flortaucipir binding in TBI participants in the right lateral occipital cortex, and this also was associated with reduced fractional anisotropy (FA) of white matter tracts.^97^ Similarly, Mohamed and colleagues (2022) reported cognitively impaired subjects with TBI had increased Flortaucipir SUVR in the superior, middle, and inferior occipital gyri compared to those with no TBI, but no differences in the lingual gyrus.^98^ In addition, TBI subject 3 from Mountz and colleagues (2017) had elevated Flortaucipir uptake in the bilateral occipital lobe,^100^ and their 2018 paper reported the three TBI patients with numerous TBIs showed high Flortaucipir uptake in the occipital lobes.^101^ Oppositely, Mohamed and colleagues (2019) saw no significant differences in Flortaucipir uptake in the lingual gyrus, fusiform cortex, or pericalcarine visual cortex of subjects with TBI compared to control subjects without TBI.^99^ Together, the majority of reports are supportive of a link between TBI history and increased occipital lobe tau. However, as not all reports or occipital ROIs demonstrated significant differences, it is important to consider that associations may be region-specific and potentially related to the number of injuries a subject experienced.

### Precuneus

Risacher and colleagues (2021) observed increased tau in the precuneus of adults with TBI with or without LOC.^95^ Similarly, Mohamed and colleagues (2019) found subjects with TBI + PTSD showed higher mean Flortaucipir SUVR in the precuneus compared to control subjects without TBI.^99^ They also found in a separate study (Mohamed and colleagues, 2022) that cognitively normal subjects with TBI had higher tau in the precuneus than those without TBI. TBI subject 3 from Mountz and colleagues (2017) also demonstrated elevated Flortaucipir uptake in the precuneus,^100^ and the three TBI patients with numerous TBIs from Mountz and colleagues (2018) exhibited similarly higher Flortaucipir uptake in the precuneus than controls.^101^ These studies further implicate the precuneus as a highly sensitive region for pathological protein deposition after TBI, in tandem with the aforementioned results from amyloid studies. The congruence of the findings speak to the need for further studies investigating why the precuneus seems to be particularly vulnerable to amyloid and tau deposition.

### Insula

Mohamed and colleagues (2019) reported significantly higher mean Flortaucipir SUVR in the insula of TBI subjects compared to control subjects without TBI.^99^ However, as no other studies examined an insular ROI, future replications are necessary to confirm these results.

### Basal ganglia

Subjects with TBI from Mohamed and colleagues (2019) exhibited significantly higher mean Flortaucipir SUVR in the basal ganglia compared to control subjects without TBI.^99^ Similarly, TBI subject 2 from Mountz and colleagues (2017) had intense Flortaucipir uptake in the left pallidum.^100^ These studies provide initial evidence that the basal ganglia may be prone to tau deposition after TBI, but as only two studies examined a basal ganglia ROI and the number of subjects was small, replications in larger cohorts are essential. Additionally, off-target binding of flortaucipir in the basal ganglia must be considered as a possible source of false positivity, and these results should be interpreted cautiously.^67^

### Substantia nigra

TBI subject 1 from Mountz and colleagues (2017) showed generally minimal Flortaucipir uptake, but slightly increased uptake in the substantia nigra relative to background levels.^100^ Similar to the insular ROI, no other studies examined the substantia nigra, so this observation warrants replication in additional subjects.

### Cingulate gyrus

Mountz and colleagues (2018) observed that TBI patients with numerous injuries exhibited high Flortaucipir uptake in the posterior cingulate gyrus.^101^ This is in line with results from studies using amyloid tracers, which found greater amyloid deposition in the cingulate; however, limited conclusions about the propensity of the cingulate to accumulate tau can be made as this is the only study to have examined tau in a cingulate ROI. As with aforementioned ROIs such as the insula and substantia nigra, additional studies are urgently needed.

## Conclusion

There are widespread but inconsistent findings in the current body of literature regarding the presence of an association between TBI and amyloid and/or tau deposition that can be visualized using PET.

In limbic regions of interest (entorhinal cortex and hippocampus), evidence for amyloid changes in individuals with TBI was mixed. Some studies reported increased amyloid in the hippocampus and entorhinal cortex, while others saw no change or even decreased amyloid. These contrasting findings were also notable in subcortical regions, including the striatum (caudate and putamen) and the cerebellum. Only one study examined patterns of amyloid deposition in the white matter, but observed subjects who had DAI showed increased amyloid tracer binding in white matter compared to patients with focal injuries. As DAI by definition causes widespread white matter damage, this finding is fairly logical; however, the potential for off-target amyloid tracer binding in the white matter representing an explanation for this observation must be carefully considered.

In cortical regions, the evidence for elevated amyloid deposition in subjects with TBI was similarly conflicted. In studies that utilized cortical summary ROIs, there was no clear consensus. While four studies found some evidence for increased global amyloid in TBI subjects, particularly in individuals with concurrent cognitive impairment, eight studies did not find associations.

On a region-specific basis, studies examining various frontal lobe ROIs were still very mixed, but most saw at least some evidence for altered amyloid deposition on a gyrus-specific level. This indicates there may be significant heterogeneity within the pattern of frontal lobe amyloid deposition. Injury severity may also be influential, as patients with DAI seemed to show consistently different patterns of amyloid deposition compared to patients with focal injuries.

Findings in the parietal lobe were slightly more clear, pointing to a general trend of altered amyloid deposition after TBI with the continued caveat of potential influences of injury severity and psychiatric comorbidities. Oppositely, there were numerous contradictions in the temporal and occipital lobes. Evidence for both increases and decreases in amyloid deposition levels were observed, and there was additional evidence that suggested no change in amyloid levels. Similarly, in the corpus callosum, one paper found no evidence for amyloid deposition while another observed increased deposition. The highly contradictory nature of the findings in many of these ROIs once again emphasizes the considerable need for replicative studies in greatly expanded samples.

Analysis of the cingulate gyrus offered slightly more clarity, with the majority of papers suggesting both the anterior and posterior cingulate might be prone to amyloid deposition in TBI cases. This also extended to the cuneus/precuneus, as all but one paper that examined this region provided supportive evidence for increased amyloid deposition in TBI subjects.

Fewer studies examined tau deposition, but the results were generally in better agreement with each other than those from amyloid studies. In the limbic regions, there was fairly consistent evidence for increased tau deposition in the medial temporal lobe/hippocampus and the temporal pole in TBI cases, whereas findings in the entorhinal cortex were mixed. In the cortical composite regions of interest, two studies noted increased tau in the mesial-temporal and temporoparietal composite ROIs in TBI subjects, but one study found no such associations. Similarly, in the neocortical composite, one study found evidence for increased tau SUVRs in TBI cases, but another did not. Finally, in the meta-temporal composite, one study found evidence for elevated tau deposition, but no other study examined this particular composite ROI to confirm or contrast these observations.

There was more consensus in the frontal lobe, where three studies all found evidence for increased tau in at least one frontal lobe region in TBI subjects. This was recapitulated in the parietal lobe, where five studies found supportive evidence for increased tau. Findings were more conflicting in the temporal and occipital lobes, as four studies in each lobe observed increased tau in TBI cases, but one in each lobe did not. Jointly, however, the bulk of these studies offer slightly improved consensus compared to observations from amyloid tracers, and suggest a general trend towards increased tau deposition in cortical lobes in subjects with TBI.

In the less studied regions of interest, one study found increased tau in the cingulate gyrus, one found increased tau in the insula, and one found increased tau in the substantia nigra. Two studies reported elevated tau in the basal ganglia, suggesting the basal ganglia may be more prone to tau deposition than amyloid deposition after TBI, as findings from amyloid studies were mixed. Though each of the aforementioned ROIs was subject to limited analysis and requires confirmatory replications, the evidence favors increased tau in subjects with TBI history.

Five studies investigated tau deposition in the precuneus, and each reported evidence for elevated tau in subjects with TBI. In line with those reports that examined amyloid deposition in this region, these findings strongly suggest the precuneus is particularly vulnerable to pathological protein deposition in subjects who have experienced a TBI.

In summary, the regions which had the strongest supportive evidence for increased amyloid deposition after TBI were the cingulate gyrus, cuneus/precuneus, and parietal lobe, while the strongest evidence for increased tau in was in the medial temporal lobe and entorhinal cortex, as well as the precuneus, frontal, temporal, parietal, and occipital lobes. Conflicting results amongst these and other ROIs reveals the great need for future studies with large sample sizes and diverse injury types. Additionally, further investigation into the impact of comorbid psychiatric diagnoses, such as PTSD, on protein deposition in subjects who have TBI history is also warranted. Consistently sparse findings from both amyloid and tau studies highlight the immediate need for replication and confirmation of findings in larger datasets in order to better characterize the association of TBI with AD-related protein accumulation on PET in aging populations.

## Transparency, Rigor, and Reproducibility Summary

No review design or strategy was preregistered for the purpose of this review. We uploaded the searches conducted for each database to searchRxiv as a search repository, and followed the PRISMA guidelines for systematic reviews to ensure reproducibility and rigor. Additionally, we took steps to ensure rigor during the screening process, including using Covidence to blind the screening process, having at least two reviewers screen each article at both steps of the screening process, and discussing conflicts in the screening process in order to come to a consensus decision for each article.

## Data Availability

All data utilized in the present study are available upon reasonable request to the authors.

## Acknowledgements

The authors thank Dr. Sujuan Gao, Dr. Karmen Yoder, Dr. Kelly Nudelman, Dr. Tom McAllister, Dr. Yu-Chien Wu, and Dr. Wei Wu for valuable discussions.

## Authorship Confirmation/Contribution Statement

KD contributed to the conception, design, literature search, screening process, and drafting of the manuscript. CV conducted the literature search and contributed to drafting of the methods section. SC and DD contributed to the screening process. SR and AS contributed to conception and revision of the manuscript. All authors read and approved the final version of the manuscript.

