## Supplemental Text for "Traumatic brain injury and Alzheimer’s Disease biomarkers: A systematic review of findings from amyloid and tau positron emission tomography (PET)"

Cochrane Database Search Terms:

Search Name: PET and Alzheimer’s

Date Run: 09/01/2023 16:00:32

Comment:

| ID | Search | Hits |
|---|---|---|
| #1 | [mh "positron emission tomography"] | 1164 |
| #2 | Positron-Emission-Tomography | 5330 |
| #3 | PET | 8554 |
| #4 | Tau-positron-emission-tomography | 6 |
| #5 | tau-PET | 56 |
| #6 | Florbetaben OR Neuraceq | 37 |
| #7 | Flortaucipir OR Tauvid | 23 |
| #8 | Flutemetamol | 33 |
| #9 | {OR #1-#8} | 9867 |
| #10 | Microtubule-associated-protein-tau | 18 |
| #11 | [mh "tau Proteins"] | 94 |
| #12 | [mh "Amyloid beta-Protein Precursor"] | 248 |
| #13 | amyloid | 2869 |
| #14 | tau-protein* | 269 |
| #15 | protein-tau* | 34 |
| #16 | tau | 6569 |
| #17 | MAPT | 149 |
| #18 | tauopath* | 65 |
| #19 | [mh "Tauopathies"] | 3975 |
| #20 | [mh "Alzheimer Disease"] | 3874 |
| #21 | alzheimer* | 14468 |
| #22 | {OR #10-#21} | 21753 |
| #23 | [mh "brain injuries"] | 2795 |

#24 brain-injur* 7993

#25 head-injur* 1979

#26 concussion* 1080

#27 neurotrauma* 456

#28 TBI 3323

#29 {OR #23-#28} 10563

#30 #9 and #22 and #29 21

Embase <1974 to 2023 January 06>

| | | |
|---|---|---|
| 1 | exp positron emission tomography/ | 214359 |
| 2 | Positron-Emission-Tomography.mp. | 227720 |
| 3 | PET.mp. | 222819 |
| 4 | Tau-positron-emission-tomography.mp. | 245 |
| 5 | tau-PET.mp. | 1433 |
| 6 | (Florbetaben or Neuraceq).mp. | 1278 |
| 7 | (Flortaucipir or Tauvid).mp. | 1488 |
| 8 | Flutemetamol.mp. | 977 |
| 9 | 1 or 2 or 3 or 4 or 5 or 6 or 7 or 8 | 312849 |
| 10 | Microtubule-associated-protein-tau.mp. | 2974 |
| 11 | exp tau protein/ | 35702 |
| 12 | exp amyloid precursor protein/ | 20721 |
| 13 | exp amyloid beta protein/ | 53318 |
| 14 | exp "amyloid beta protein[1-40]"/ | 6966 |
| 15 | exp amyloid protein/ | 5521 |
| 16 | exp amyloid beta protein antibody/ | 928 |
| 17 | exp "amyloid beta protein[25-35]"/ | 1894 |
| 18 | exp "amyloid beta protein[1-42]"/ | 16866 |
| 19 | amyloid.mp. | 158997 |
| 20 | tau-protein*.mp. | 37872 |
| 21 | protein-tau*.mp. | 4200 |
| 22 | tau.mp. | 78529 |
| 23 | MAPT.mp. | 3768 |
| 24 | tauopath*.mp. | 9606 |
| 25 | exp tauopathy/ | 7168 |
| 26 | exp Alzheimer Disease/ | 236017 |
| 27 | alzheimer*.mp. | 290465 |

28 10 or 11 or 12 or 13 or 14 or 15 or 16 or 17 or 18 or 19 or 20 or 21 or 22 or 23 or 24 or 25 or 26 or 27 395884

29 exp brain injury/ 205692

30 brain-injur*.mp. 182866

31 head-injur*.mp.62960

32 concussion*.mp. 19590

33 neurotrauma*.mp. 4243

34 TBI.mp. 50163

35 29 or 30 or 31 or 32 or 33 or 34 290877

36 9 and 28 and 35 803

Ovid MEDLINE(R) ALL <1946 to January 06, 2023>

| | | |
|---|---|---|
| 1 | exp Positron-Emission Tomography/ | 76608 |
| 2 | Positron-Emission-Tomography.mp. | 112687 |
| 3 | PET.mp. | 123875 |
| 4 | Tau-positron-emission-tomography.mp. | 194 |
| 5 | tau-PET.mp. | 674 |
| 6 | (Florbetaben or Neuraceq).mp. | 385 |
| 7 | (Flortaucipir or Tauvid).mp. | 319 |
| 8 | Flutemetamol.mp. | 296 |
| 9 | 1 or 2 or 3 or 4 or 5 or 6 or 7 or 8 | 157080 |
| 10 | Microtubule-associated-protein-tau.mp. | 2287 |
| 11 | exp tau Proteins/ | 18048 |
| 12 | exp Amyloid beta-Protein Precursor/ | 46667 |
| 13 | amyloid.mp. | 114133 |
| 14 | tau-protein*.mp. | 21268 |
| 15 | protein-tau*.mp. | 3188 |
| 16 | tau.mp. | 55395 |
| 17 | MAPT.mp. | 4646 |
| 18 | tauopath*.mp. | 5545 |
| 19 | exp Tauopathies/ | 118509 |
| 20 | exp Alzheimer Disease/ | 114419 |
| 21 | alzheimer*.mp. | 193967 |
| 22 | 10 or 11 or 12 or 13 or 14 or 15 or 16 or 17 or 18 or 19 or 20 or 21 | 278586 |
| 23 | exp Brain Injuries/ | 80268 |
| 24 | brain-injur*.mp. | 108612 |
| 25 | head-injur*.mp. | 29387 |
| 26 | concussion*.mp. | 16303 |
| 27 | neurotrauma*.mp. | 2615 |

| | | |
|---|---|---|
| 28 | TBI.mp. | 30310 |
| 29 | 23 or 24 or 25 or 26 or 27 or 28 | 143599 |
| 30 | 9 and 22 and 29 | 172 |
